# A STUDY OF FACTORS INFLUENCING FETAL MONITOR FAILURE

**DOI:** 10.64898/2026.07.12.26357884

**Authors:** E Enkh-Urel, B Odgerel, S Ichinkhorloo, T Altangerel, T Amgalan, B Saranzaya, Ts Dashdulam

**Affiliations:** Department of Physics and Informatics, School of Biomedicine, Mongolian National University of Medical Sciences, Ulaanbaatar, Mongolia; Department of Basic Science, School of Nursing, Mongolian National University of Medical Sciences, Ulaanbaatar, Mongolia; Department of Basic Medical Sciences, School of Nursing, Mongolian National University of Medical Sciences, Ulaanbaatar, Mongolia

**Keywords:** Fetal Monitoring, Equipment Failure, Risk Factors

## Abstract

In Mongolia, an average of 65,000 women become pregnant each year, and about 59,500 babies are born. Although the number of pregnancies is decreasing by 8–12 percent each year, the level of fetal monitor usage remains high. The capital’s maternity hospital currently has 27 fetal monitors in use, and an average of 30–35 calls are recorded per month. However, there is a lack of research on the use of fetal monitors, the causes and influencing factors of damage, and the organization of technical services. Therefore, this topic was chosen to determine the usage status of fetal monitors, the causes of malfunctions, and ways to improve them.

**Purpose:** To study the causes and factors affecting possible damage and injury during the use of fetal monitors, and to identify ways to reduce them.

**Materials and methods:** A one-time study was conducted on 10 MT-610 fetal monitors that were put into operation in 2019 at the Urgo Maternity Hospital in the capital. Data were collected and processed using document analysis methods from the technical passports and call logs of these devices. The factors contributing to common failures were identified using focus group interviews with the engineers and technicians responsible for the equipment.

**Results:** This study found that fetal monitor failures are caused by improper use, lack of regular calibration, electrical fluctuations, ambient temperature and humidity, and insufficient medical staff skills, training, and knowledge of how to use the device, all of which contribute to failures and measurement errors. It is also observed that when a replacement part is needed for a monitor that frequently breaks, the monitor is more likely to break again if it is used as a replacement from a previously broken monitor. Therefore, training doctors and nurses who replace spare parts on their use has been observed to significantly reduce future breakdowns.

**Conclusion:** According to the study results, the breakdowns and failures of fetal monitoring devices are mainly related to internal system failures, unstable power supply, and wear and tear of accessories and mechanical parts. The highest percentage of device failures indicates the need for special attention to the reliability of the device’s basic functions.

Additionally, the high percentage of accessory and printer failures indicates the need for proper use and monitoring of the entire device. In addition to technical factors, human misuse, lack of maintenance, and environmental influences also play a significant role in damage. Therefore, it is concluded that to ensure the reliable operation of fetal monitors, it is necessary to perform regular maintenance, stabilize the power supply, improve the quality of accessories, and increase the knowledge and skills of medical staff.

## INTRODUCTION

### Technical and Signal Extraction Pitfalls

External monitors rely heavily on ultrasound Doppler technology or maternal abdominal electrodes, both of which face severe signal extraction limitations:

- **Signal Ambiguity and Maternal Cross-talk:** One of the most critical safety issues is **Maternal Heart Rate Artefact (MHRA)**. This occurs when the ultrasound transducer accidentally locks onto the maternal abdominal aorta or uterine arteries. Modern cardiotocography (CTG) monitors use *autocorrelation algorithms* to smooth out traces, which can seamlessly transition from the fetal heart rate to the maternal pulse without creating an obvious break on the paper trace. This can dangerously mask fetal distress or even pre-existing fetal demise^13^.
- **Signal Frequency Doubling and Halving:** External monitors can experience signal processing errors where they mathematically double a low fetal heart rate (e.g., mistaking a dangerous bradycardia of 60 bpm for a normal 120 bpm) or halve a tachycardic rate, generating false alarms^14^.
- **Low Signal-to-Noise Ratio (SNR):** Non-invasive abdominal fetal electrocardiograms (fECG) struggle because the tiny electrical signal from the fetal heart must pass through layers of amniotic fluid, uterine muscle, and maternal skin. It frequently gets drowned out by maternal muscle movements (myographic signals), fetal brain activity, and transient electronic noise^15^,

### Mitigations Used in Modern Practice

To counteract these vulnerabilities, obstetric units utilize several secondary strategies:

- **Internal Monitoring:** When external traces fail, clinicians may switch to an invasive **Fetal Scalp Electrode (FSE)**, which provides direct, high-accuracy internal ECG readings unaffected by maternal BMI or movement^14^.
- **Dual-Rate Verification:** Many modern systems simultaneously track the maternal pulse via a maternal pulse oximeter alongside the fetal trace to send an alert if “signal ambiguity” (overlapping heart rates) is detected^13^.

### Equipment-Related Factors

Equipment malfunction is one of the leading causes of fetal monitor failure.

Common hardware-related problems include:

- Aging electronic components
- Ultrasound transducer failure
- Tocodynamometer malfunction
- Damaged cables or connectors
- Display and printer failures
- Battery deterioration
- Software or firmware faults

These failures can lead to inaccurate fetal heart rate (FHR) recordings, signal interruption, and device downtime, compromising continuous fetal surveillance. Studies of electronic fetal monitoring systems have highlighted that reliable equipment performance and regular servicing are essential for maintaining monitoring quality^16^.

### Sensor and Signal Acquisition Problems

The quality of CTG recordings depends on effective signal acquisition. Signal loss is frequently associated with:

- Incorrect placement of the ultrasound transducer
- Loose abdominal belts
- Damaged probes
- Insufficient ultrasound gel
- Poor skin contact
- Maternal position changes

Poor signal acquisition may cause intermittent fetal heart rate recordings, false alarms, or confusion between maternal and fetal heart rates. Maintaining proper transducer positioning throughout labor is therefore essential^17^.

### Research Gap

Although numerous studies have investigated CTG interpretation and clinical decision-making, relatively few have specifically examined the determinants of **technical fetal monitor failure**, particularly in low- and middle-income countries. Existing research indicates that monitor failure is multifactorial and that preventive maintenance, biomedical engineering support, operator competency, and organizational systems should all be considered when evaluating device reliability. Future research should integrate engineering, clinical, and human factors perspectives to develop comprehensive strategies for reducing fetal monitor failure and improving maternal–fetal safety^18^.

#### Purpose

To study the causes and factors affecting possible damage and injury during the use of fetal monitors, and to identify ways to reduce them.

#### Importance

The results of this study indicate the need to shift the policy of operation and maintenance of medical equipment from a “fix it when it breaks” approach to a “preventive risk reduction” approach. Since fetal monitors are important devices that affect maternal and fetal safety, it is important to detect their damage early, provide standardized instructions to department users, and create a mechanism for discussing the causes of repeated damage between departments. This study also highlights the need to link the work of equipment engineers and technicians to the organization’s quality management, risk assessment, budget planning, and training systems.

For example, in situations where configuration loss is a great concern, it may be more effective to include a user manual in the device rather than just replacing the battery.

#### Methodology

This work is an observational study that aggregates and evaluates records of medical equipment usage, maintenance, and damage. The study used a combination of quantitative and qualitative methods. The quantitative part used call logs, technical operation passports, breakdown repair records, and annual workload data.

The qualitative section included focus group interviews with engineers and technicians, experience notes, and an analysis of practical difficulties.

#### Scope of the study

The study was conducted based on fetal monitors used at the Urgo Maternity Hospital in the capital. A total of 27 fetal monitors of 6 types are used at the Urgo Maternity Hospital in the capital, including Bionet FC-700, Toitu MT-610, Contec CMS800G, Bionecs BFM900, F2 Edan, and FC-1400, of which 10 MT-610 devices were studied for a more detailed one-time evaluation.

**Table 1.**
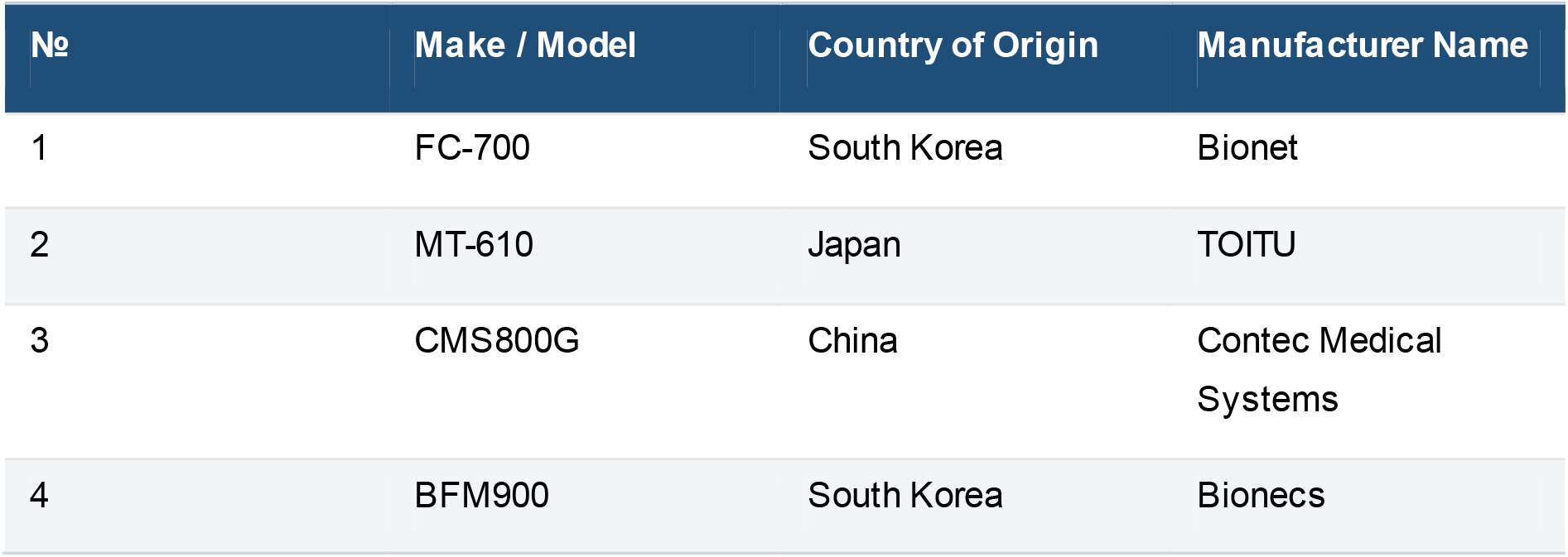
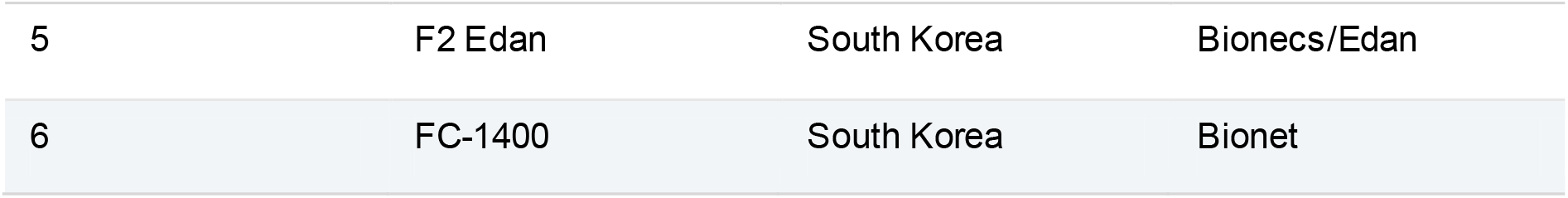
Types of fetal monitors utilized at Capital City Urgoo Maternity Hospital.

#### Research design

The research design is a cross-sectional, documentary-based, and descriptive study.

1. The structure of common failures was determined by compiling actual technical usage data.
2. Factors that may contribute to the breakdown were identified through focus group interviews.
3. The results were discussed in comparison with the review of the literature and practical needs.

#### Methodology

The research data were collected using questionnaires, interviews, documents, letters, and observation methods, including centralized engineer and technician passports and call logs.

##### Interview method

Information materials, such as fetal monitor maintenance, unexpected breakdowns, and doctors’ and nurses’ ability to communicate with fetal monitors, were developed by engineers and technicians based on the principle of creating conversations around specific topics.

##### Technical passport

It is a document that engineers and technicians record and maintain, including information such as the manufacturer, serial number, name, brand, year of manufacture, and date of commissioning of the fetal monitor, installed unit, overhaul and routine maintenance schedule, technical specifications, accessories, etc., along with inspection and maintenance frequency.

**Table 2.**
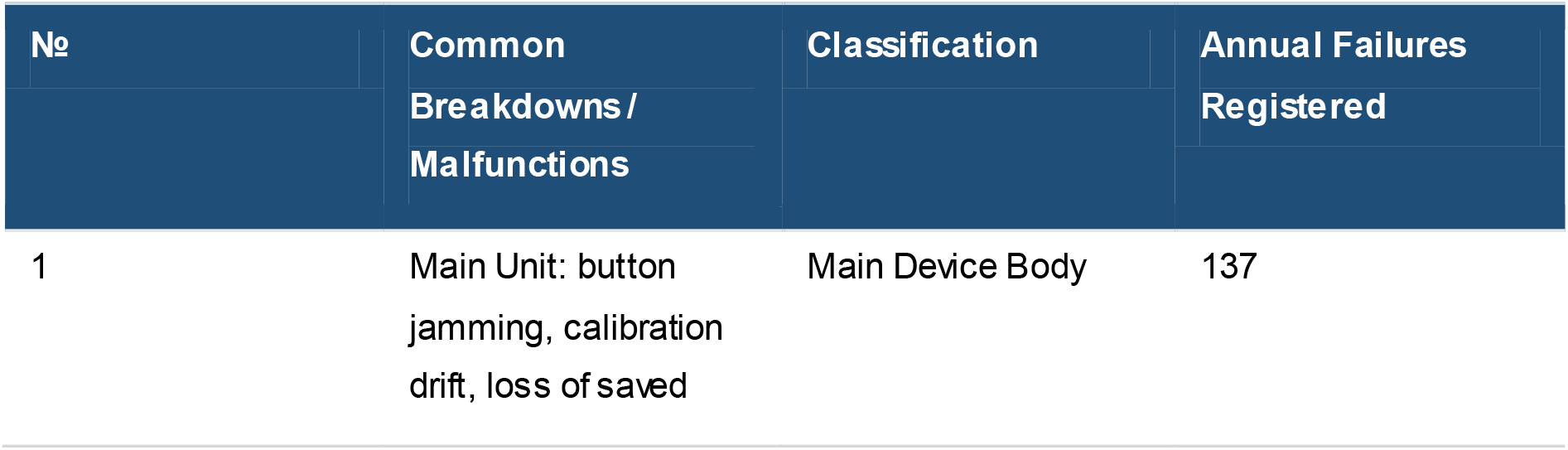
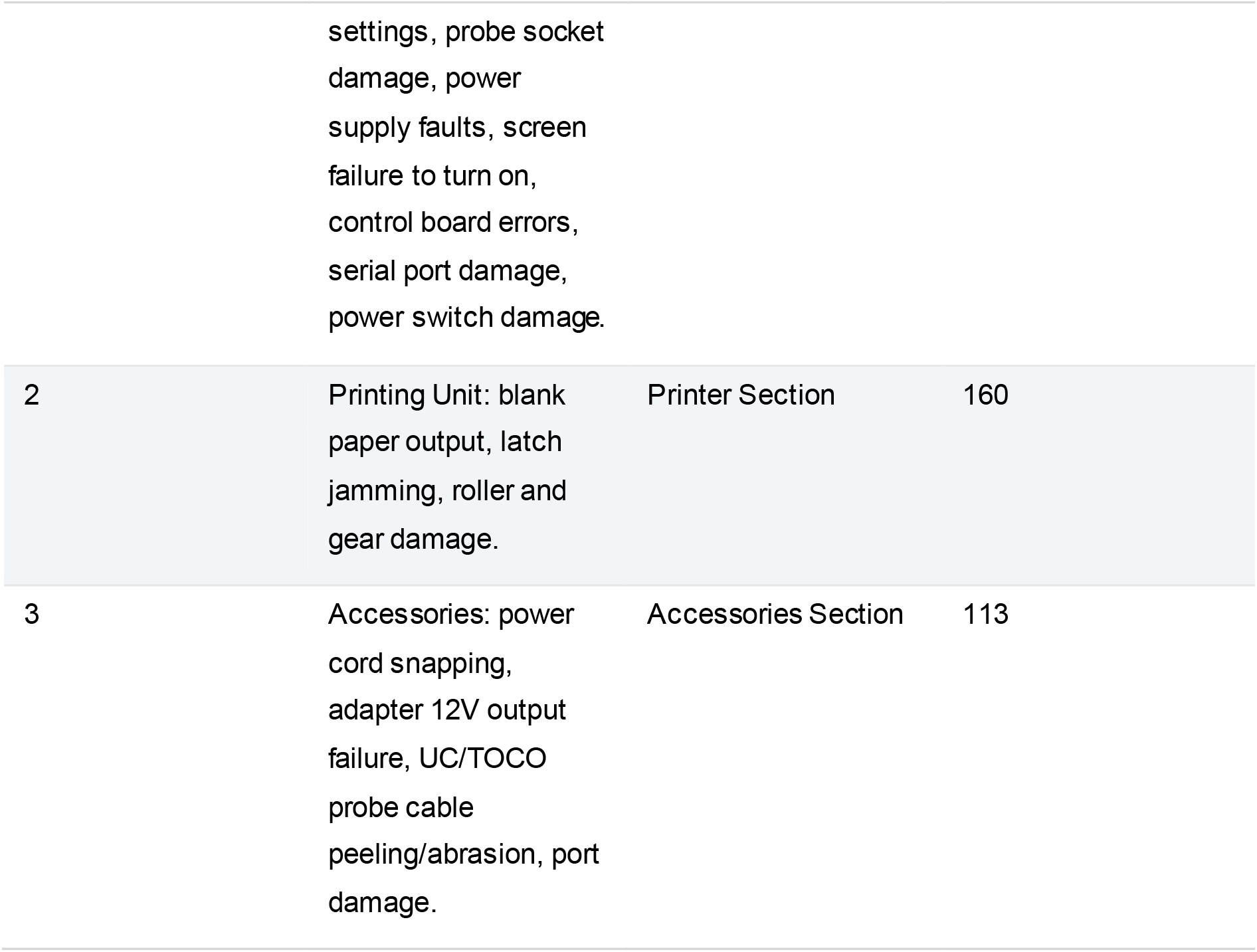
Classification of commonly occurring breakdowns and malfunctions.

| No | Common Breakdowns / Malfunctions | Classification | Annual Failures Registered |
| --- | --- | --- | --- |
| 1 | Main Unit: button jamming, calibration drift, loss of saved settings, probe socket<br>damage, power<br>supply faults, screen<br>failure to turn on,<br>control board errors,<br>serial port damage,<br>power switch damage. | Main Device Body | 137 |
| 2 | Printing Unit: blank<br>paper output, latch<br>jamming, roller and<br>gear damage. | Printer Section | 160 |
| 3 | Accessories: power<br>cord snapping,<br>adapter 12V output<br>failure, UC/TOCO<br>probe cable<br>peeling/abrasion, port<br>damage. | Accessories Section | 113 |

#### Materials and tools

The following documents and materials were used in the study: technical passports for fetal monitors, call logs, engineering commission notes, maintenance lists, and departmental equipment counts and load records.

#### Research variables and evaluation criteria

The main variables of the study are device type, department location, daily operating hours, annual load, type of failure, cause, repair method, frequency of maintenance, user training, and environmental conditions. The damage was classified into “primary,” “secondary,” and “other,” and the causes of each were identified. Influencing factors were assessed by grouping them into internal and external causes. The study did not use any personal patient information, diagnosis, or confidential treatment information, and only analyzed equipment records, technical notes, and institutional usage data. However, since internal information of the organization is being used, the principle of respecting the confidentiality of information, the reputation of the organization, and the reputation of the manufacturer was adhered to.

**Table 3.** Device counts and operational workload by participating departments. ***Note:*** As can be seen from the table above, the highest number of devices used in the delivery room and the highest daily operating hours are the reasons why the load is concentrated in this area, and the risk of breakdowns is increased.

| Department Name | Number of Devices | Available Hours / Day | Actual Operating Hours / Day | Annual Workload (Hours) |
| --- | --- | --- | --- | --- |
| Outpatient Clinic | 6 | 18 | 8 | 12,528 |
| Delivery Ward | 10 | 18 | 18 | 46,980 |
| ICU / Delivery Suite | 1 | 18 | 9 | 2,349 |
| Admissions / Triage | 2 | 18 | 18 | 2,088 |
| High-Risk Pregnancy | 1 | 18 | 8 | 12,789 |
| NICU / Emer. Delivery | 7 | 18 | 7 | 2,088 |

## Results

### Fetal monitor usage status

The use of fetal monitors at the Urgo Maternity Hospital in the capital city varied by department, but was generally high overall. Daily use is continuous in the maternity ward, high-risk prenatal care unit, and outpatient clinic. During high workloads, the same device is used on multiple machines, probes and cables are moved frequently, and the printer is constantly running, which creates conditions for accelerated mechanical wear. According to the records used in the study, the annual workload of the apparatus, which was active for up to 18 hours a day, was highest in the maternity ward. This is due to the need for continuous monitoring during childbirth. In high-traffic areas, button wear, cable damage, printer failures, and the need for reconfiguration tend to be more frequent.

#### 1. Accessory damage

This accounts for the majority of all injuries and is usually caused by mechanical factors.

- Sensor damage: The crystals inside the fetal heart rate (FHR) and TOCO sensors have cracked and lost their sensitivity.
- Cable break: The wire connecting the sensor to the device may be bent, broken, or the connector may not be making a connection.
- Fastening belt: The belt designed to hold the pressure sensor has stretched and is no longer functional.

#### 2. Damage to the body of the device

There is a defect in the device’s main processing unit and electronics.

- Mainboard: Processor and microcircuit performance may slow down, and the system may not boot.
- Screen: Scratches, discoloration, or loss of touchscreen functionality on the screen.
- Power supply unit: The device may not turn on, the battery may not hold a charge, or the fuse may blow due to sudden voltage fluctuations.

#### 3. Printer damage

- Thermal Head: Blurry graphics and white lines on the paper may occur due to a dirty or expired print head.
- Mechanical transmission: The paper feed gears may be stuck, or the paper sensor may not work.

1. According to the results of the study, the highest percentage of device failures was 44.01%. This indicates that problems related to the basic operation of the device (buttons stuck, lost settings, sensor input damage, screen malfunctions, etc.) are most common.
2. Accessory defects account for 28.39%, including power plugs, adapters, UC/TOCO probe wires, and input defects. This indicates that issues related to the quality and usability of external connections and accessories are relatively common.
3. Printer failures account for 27.60%, including issues such as blank paper, jammed latches, and damaged rollers and gears. This indicates that printing unit failures are also somewhat common.

The study shows that the highest rate of damage to the device’s main body indicates the need to pay attention to the reliable operation of the device’s main system. Additionally, a significant percentage of accessory and printer failures also occur, indicating the need to improve the comprehensive use, maintenance, and quality control of fetal monitors.

**Table 4.** Examples of malfunctions, root causes, and common repair methods. *Explanation:* As seen from the tables, many repair forms are repetitive, involving replacement, cleaning, adjustment, and connection restoration. This demonstrates that if preventative maintenance is well-organized, many malfunctions can be mitigated at an early stage.

| Malfunction Example | Root Cause | Common Repair Method |
| --- | --- | --- |
| Stuck / Jammed Buttons | Excessive pressing force, high workload, contamination, dust accumulation | Clean button assembly, replace/refurbish if necessary |
| Calibration / Setting Loss | User changes, incorrect base settings modification, low internal battery | Restore standard configurations, replace 3V battery, conduct user training |
| Blank Paper Output | Contamination of print path, incorrect settings, faulty print head | Clean the print path, verify configuration, inspect/service thermal head |
| Probe Socket Damage | Forceful plugging/unplugging, twisting cables, dropping components | Repair socket connection, re-solder, replace connector |
| Adapter Output Failure | Power grid fluctuations, adapter aging | Test adapter output, replace with a new unit |
| UC/TOCO Cable Peeling | Frequent transport, severe cable bending, outer sheath wear | Apply cable protection, replace assembly if necessary |

#### Factors affecting injuries

According to the focus group interviews and registration results, human error was identified as the most influential factor in the damage. This includes pulling the probe hard, bending the cable, abruptly closing the printer lid, accidentally changing settings, connecting without completely drying it after cleaning, and roughly unplugging the power plug. In interviews with engineers and technicians, the idea that “most small failures start with mishandling” was repeated many times.

Environmental influences include improper storage location, humidity, dust, poorly ventilated rooms, electrical fluctuations, and lack of an uninterrupted power source. In particular, power fluctuations may negatively affect the lifespan of the adapter, main power module, and internal battery. Also, in some departments, the mechanical stress on the probe and cable is increasing due to the constant movement of the device.

Long-term stress causes the device to operate continuously during peak labor, the printer to record continuously, buttons to be pressed repeatedly, and the probe to be moved frequently. Internal factors include component aging, battery capacity decline, sensor wear, software failures, and loose connector contacts and oxidation. Therefore, it is necessary to consider multiple system factors together rather than considering a failure solely as a result of user error.

**Table 5.** Distribution of factors influencing equipment breakdowns.

| No | Influencing Factors | Frequency (Score) | Percentage (%) |
| --- | --- | --- | --- |
| 1 | User-handling errors /<br>Improper operation | 21 | 65.6% |
| 2 | Long-term operational<br>workload stress | 6 | 18.8% |
| 3 | Environmental<br>influences / Grid<br>instability | 5 | 15.6% |

#### Technical inspection service status

According to the technical inspection service records, scheduled inspections are performed regularly every month, but the number of unscheduled services performed when a breakdown occurs is several times higher. This indicates that the operating conditions of the device are severe, or that all risks cannot be fully mitigated within the framework of a scheduled inspection. While preliminary inspection services were conducted quarterly and routine inspections were conducted daily, it is positive that disruptions related to user habits and load continue.

**Table 6.**
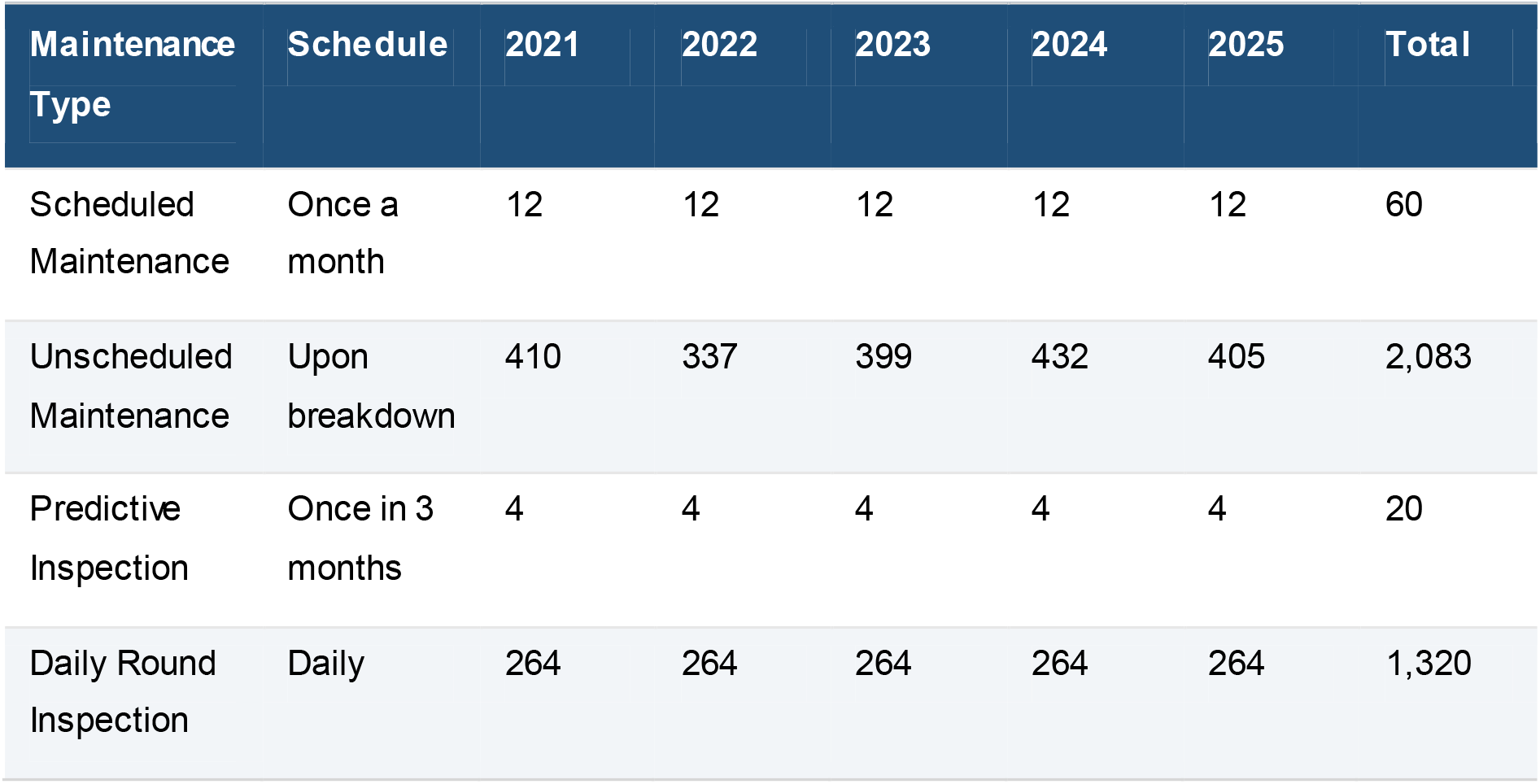
Frequency of technical inspection and maintenance tasks.

| Maintenance Type | Schedule | 2021 | 2022 | 2023 | 2024 | 2025 | Total |
| --- | --- | --- | --- | --- | --- | --- | --- |
| Scheduled Maintenance | Once a month | 12 | 12 | 12 | 12 | 12 | 60 |
| Unscheduled Maintenance | Upon breakdown | 410 | 337 | 399 | 432 | 405 | 2,083 |
| Predictive Inspection | Once in 3 months | 4 | 4 | 4 | 4 | 4 | 20 |
| Daily Round Inspection | Daily | 264 | 264 | 264 | 264 | 264 | 1,320 |

**Table 7.** Detailed annual frequency of common malfunctions.

| No | Common Breakdowns / Malfunctions | Annual Total Count |
| --- | --- | --- |
| 1 | Configuration / Setting Loss | 100 |
| 2 | Blank Paper Output | 60 |
| 3 | Latch Jamming or Damage | 50 |
| 4 | Roller and Gear Breakdowns | 50 |
| 5 | Button Jamming / Sticking | 25 |
| 6 | Power Plug 220V Snapping / Abrasion | 42 |

This table shows that the highest frequency of configuration loss issues is related to user training, standard settings, and internal battery status. White paper jams and roller damage indicated that the printer was overloaded and mechanically worn. Sensor and adapter issues also accounted for a high percentage, highlighting the need to improve the protection, storage, and transportation of accessories.

**Table 8.** Primary conclusions drawn from technical focus group sessions.

| Problem Dimension | Key Observation / Focus Group Conclusion |
| --- | --- |
| Most Vulnerable Components | Printer covers, locking latches, feed rollers, probe cables, and input sockets. |
| Primary Root Causes | High operating workloads, improper user habits, acute cable bending, and power line voltage spikes. |
| Repair Bottlenecks | Procurement delays for specific spare parts; highly restricted availability of temporary backup units. |
| Preventative Methods | Routine multi-point inspections, structured cleaning, brief user training, and standardized inter-ward protocols. |
| Budgetary Implications | Frequent minor breakdowns significantly inflate aggregate expenditure; strategic spare parts inventory planning is critical. |

The interviews show that many issues considered “minor breakdowns” continue to occur in reality, increasing the overall maintenance burden. Experts believe that simple measures such as brief user training, proper probe and cable storage, standard operating procedures, and power protection are effective in reducing repetitive strain injuries.

## Discussion

### Causes and effects of trauma

The results of the study showed that fetal monitor failures are caused by a combination of factors. The most frequently encountered configuration loss problem is not only due to electronic failures, but can also be due to frequent changes in the user interface, non-compliance with standard procedures, and a weak internal memory or battery. While this type of disruption may not seem to directly impact clinical decisions, the quality of monitoring can be compromised if recording speed, timing, patient information, and alarm thresholds are incorrect. The high frequency of printing area damage is related to the specific use of fetal monitors. Since real-time recording is common in prenatal care, the paper, rollers, and thermal printer tracks and latches are subject to significant mechanical stress. Therefore, improper user behavior, such as closing the lid forcefully, using the wrong paper, and not cleaning it on time, can lead to damage. Accessory damage, such as frayed probe wires, loss of adapter output, and damage to input connectors, is related to transporting the device, unprotected cable wrapping, and unstable power lines. Therefore, damage is not just a manifestation of the age of the device, but also an indicator of the management of the operating environment.

### Comparison with other research and practice

A common trend in the use of medical equipment is that mechanical wear, cable damage, power problems, and misuse are prevalent in high-traffic wards. In the case of fetal monitors, the probe and printer are parts that are directly involved with the user, so repeated failures in these are common in international practice. The results of this study also confirm the same trend. In other words, it is not the age of the device alone that determines the failure, but rather the failure of the three-way relationship: “user-device-environment”. Experience with other medical device maintenance, such as ultrasound machines, patient monitors, and infusion pumps, shows that poor user practices, lack of ongoing training, and lack of advance spare parts planning can turn minor breakdowns into systemic problems. Fetal monitor results are also consistent with this conclusion, so an organization-wide equipment education program is needed.

## Conclusion

According to the results of the study, failures and damage to fetal monitoring devices are usually related to wear and tear on the device’s internal systems, power supply, accessories, and mechanical components. In particular, the highest rate of damage to the device’s main body indicates the need for special attention to the reliability of the device’s basic functions. Additionally, a significant percentage of equipment and printer failures are also occurring, indicating the need for proper use and monitoring of all equipment.

In addition to technical factors, it has been found that human misuse, lack of maintenance, and environmental influences also play an important role in the occurrence of breakdowns. Therefore, it is concluded that to ensure the reliable operation of fetal monitors, it is necessary to perform regular maintenance, stabilize the power supply, improve the quality of accessories, and increase the knowledge and skills of medical staff.

## Research limitations

The study used data from a single institution, a specific time period, and focused evaluation on the MT-610 device, which limits the direct generalization of the results to hospitals at all levels. Also, some records were kept manually, the accuracy of damage descriptions varied, and information on repair times and costs was incomplete.

Focus group interviews have practical value, but since they are qualitative data, they are also dependent on the experiences of the participants. However, the strength of this work is that it combines actual usage records, engineering observations, and departmental load data to form an assessment. Further research could include studies that include multiple hospitals, compare long-term trends, and consider item-level reliability.

## Data Availability

Data is available.

